# Educational Impact of a Research and Mentoring Symposium Emphasizing Formative Feedback for Medical Students and Early Career Doctors in Africa

**DOI:** 10.1101/2023.11.03.23298036

**Authors:** Anya A Nowbuth, Mwiza Muwowo, Mwitupa Makashinyi, Andrew Kumwenda, Sheila J Mwanamwampula, Tamara Kaluba, Sula Mazimba, Seth M Bloom, Akwi W Asombang

## Abstract

Research training is a core component of medical education, but many African medical schools lack resources to support student research, impeding global health progress. Conferences offer alternate venues to learn about research, network, and receive scientific feedback, but conference access for African trainees is limited. We hypothesized that a research and mentorship symposium for African medical trainees could promote research knowledge and interest among attendees.

**Methods:** We co-organized a symposium attended by medical students and early career doctors from African institutions in-person (Lusaka, Zambia) and virtually. The program featured trainee abstract presentations, keynote lectures, and networking. Abstracts received written reviews and judges provided live formative feedback on presentations. Participants completed a post-symposium survey on demographics, institutional research support, and benefits of symposium participation.

**Results:** Respondents included 87 trainees from 7 African countries, including 28 presenters from 11 schools. Most had never attended or presented at a conference, interacted with peers in a research forum, or received formal research training. The majority reported key unmet needs in research training and resources at their institutions. Trainees praised the symposium’s judging format and said attendance inspired them, increased their interest in research, improved quality of their projects, and motivated them to initiate new studies.

**Interpretation:** A volunteer-organized research and mentorship symposium emphasizing formative feedback enhanced research knowledge and interest among African medical trainees, many of whom had limited access to institutional research training and support. Such initiatives can inspire and nurture new generations of African scientists to advance global health.

**Funding:** None

## Background

Training in the conduct and interpretation of research is a core component of medical education and essential for developing a robust global health workforce. World Federation for Medical Education Global Standards for Quality Improvement recommends incorporating principles of the scientific method, analytical and critical thinking, medical research methods, and evidence-based medicine throughout medical school curricula, and encourage original research as part of training.^1,2^ Research experiences improve students’ comfort understanding research, increase their likelihood of conducting research in future, influence specialty choice, and increase odds of pursuing academic careers.^3–6^ However, many African medical schools lack robust research training infrastructure and opportunities^7–9^, which can be associated with greater likelihood of research-inclined medical trainees emigrating to high-income countries.^10^ The Sub-Saharan African Medical School Study (SSAMSS) of >100 African institutions found that <10% of faculty members at most schools were involved in sponsored research, limiting knowledge generation, impeding recruitment and retention of talented faculty members, and harming student training.^11^ SSAMSS researchers recommended improved funding for research and research training at African institutions to improve medical education and population health, while emphasizing need for creative solutions and international collaborations to overcome scarce institutional support.

Professional and trainee-organized conferences offer alternate venues for trainees to learn about research, develop scientific networks, and gain feedback on research and presentation skills.^12^ Conferences furnish opportunities to meet external collaborators and mentors, which holds particular value for trainees lacking institutional research support.^13^ Trainee-targeted conferences are broadly available in high-income countries – one study identified 27 student/trainee medical conferences in a 4-month period in the United Kingdom alone.^14^ Systematic data on African conferences are lacking, but conference access is anecdotally much more limited for African trainees.

We hypothesized that a medical research and mentoring symposium for African trainees could help address this need by promoting research knowledge and enthusiasm among attendees. With volunteers from multiple organizations, we co-organized a hybrid in-person/virtual symposium in Lusaka, Zambia, for students and recent graduates from diverse African medical schools. The program featured keynote lectures and trainee abstract presentations with emphasis on live, public formative feedback for trainees from judges. Attendees completed a survey on demographics, prior research experience, institutional research support, and benefits from symposium attendance. Most reported major unmet needs in medical school research training and support, while endorsing substantial benefits from symposium attendance. These results highlight the global health workforce impacts achievable through such an initiative.

## Methods

### Conference Format

The symposium occurred on 18-19 October, 2022, at the Paediatric Centre of Excellence - University Teaching Hospitals in Lusaka, Zambia. Virtual attendees participated via Zoom. It was co-organized by the Pan-African Organization for Health Education and Research (POHER), the Copperbelt University School of Medicine (CBU-SOM) Mentorship Program, Young Emerging Scientists Zambia (YES Zambia), and Women in Nuclear Zambia (WiN Zambia). This was the fourth annual symposium, but recent prior editions were entirely virtual due to the COVID-19 pandemic. Organizers were unpaid volunteers, primarily comprising current students or recent graduates of African medical schools. The program featured trainee oral abstract presentations, as well as keynote addresses by speakers from Zambia, Kenya, the United States of America, Canada, and Dominica on topics including research, policy, mentorship, and trainee mental health. The program also included networking, remarks from judges, and a presenter awards ceremony (Supplementary Table ST1).

Research and clinical case report abstracts from students and recent (within two years) graduates of African medical schools were scored by an international panel of reviewers blinded to authorship, who also provided written formative feedback. Abstract presenters gave 7-minute presentations, followed by 5-minute question-and-answer sessions that included audience questions and live formative feedback from senior judges (S.M.B. and A.K.). Prizes consisting of certificates and monetary awards were given for top abstracts and presentations. “People’s choice” prizes were assigned based on audience voting. The total prize budget was 750 US dollars (USD), divided among awardees.

### Data Collection

Conference registrants received an electronic Qualtrics-based survey, which was distributed after the final abstract presentation and remained open for 12 days. Eligibility to vote for “people’s choice” prizes and receive symposium certificates of attendance was conditional on survey completion. Abstract presenters completed a supplementary survey on their experience presenting.

### Analysis

Responses were exported as Microsoft Excel files. Data cleaning, analysis, and plotting was performed in R version 4.2.1^15^ using *tidyverse* package version 2.0.0.^15,16^ We excluded three duplicate responses, a clearly spurious response, and incomplete surveys that failed to answer any of the questions used for analysis. Respondents were classified as “trainees” in the analysis if they were currently enrolled medical or health sciences students or junior doctors (interns). We defined five categories of trainee satisfaction with institutional research support. Trainees were considered satisfied in these categories if they replied “somewhat agree” or strongly agree” to questions about 1) whether their school encourages students to pursue research as part of training, 2) whether their school provides research opportunities, 3) whether their school provides adequate training in research, 4) whether students at their school were satisfied with access to research training and mentorship, or 5) whether the respondents individually had adequate access to research mentorship. We also defined six categories of benefit from the symposium. Respondents were classified as benefiting if they replied “somewhat agree” or strongly agree” to questions about 1) whether the symposium increased their interest in research, 2) whether research topics were informative and educational, 3) whether listening to others present was inspirational, 4) whether judge feedback inspired their own research, or 5) if the symposium “somewhat” or “extremely” increased their likelihood of submitting an abstract to another conference, or 6) if they derived “good”, “very good”, or “excellent” benefit from keynote presentations. Respondents failing to answer any of the latter six questions were excluded from benefit analysis, while those who answered at least one were classified as not benefitting in the omitted categories.

Free-response answers about which aspects participants liked, disliked, or would change about the symposium and judging were thematically analyzed. Common response elements were grouped into themes and further categorized into subthemes until no new themes were evident.

### Ethical Approval

Informed consent was obtained prior to voluntary survey participation. Ethics approval was obtained from the Tropical Diseases Research Center (TDRC) Ethics Committee [TDREC/023/022] in Ndola, Zambia.

## Results

### Attendee Characteristics

Ninety-four valid survey responses were included in the analysis, representing a response rate of 65·6% among 154 total registrants. However, this underestimates the true attendee response rate, since not all registrants attended and the hybrid format prevented determining precise attendance. Respondents included 87 trainees and 7 non-trainees (table 1). Most trainees (n=76, 87·4%) were current medical students and a slight majority (n=44, 50·6%) reported their gender as female. The largest number attended from Zambia (n=72, 82·8%), followed by Egypt (n=6, 6·9%), Kenya (n=4, 4·6%), and Nigeria (n=2, 2·3%). One trainee each attended from South Africa, Cameroon, and Sudan (figure 1A, table 1). Sixty-one (70·1%) attended in-person.

**Figure 1.**
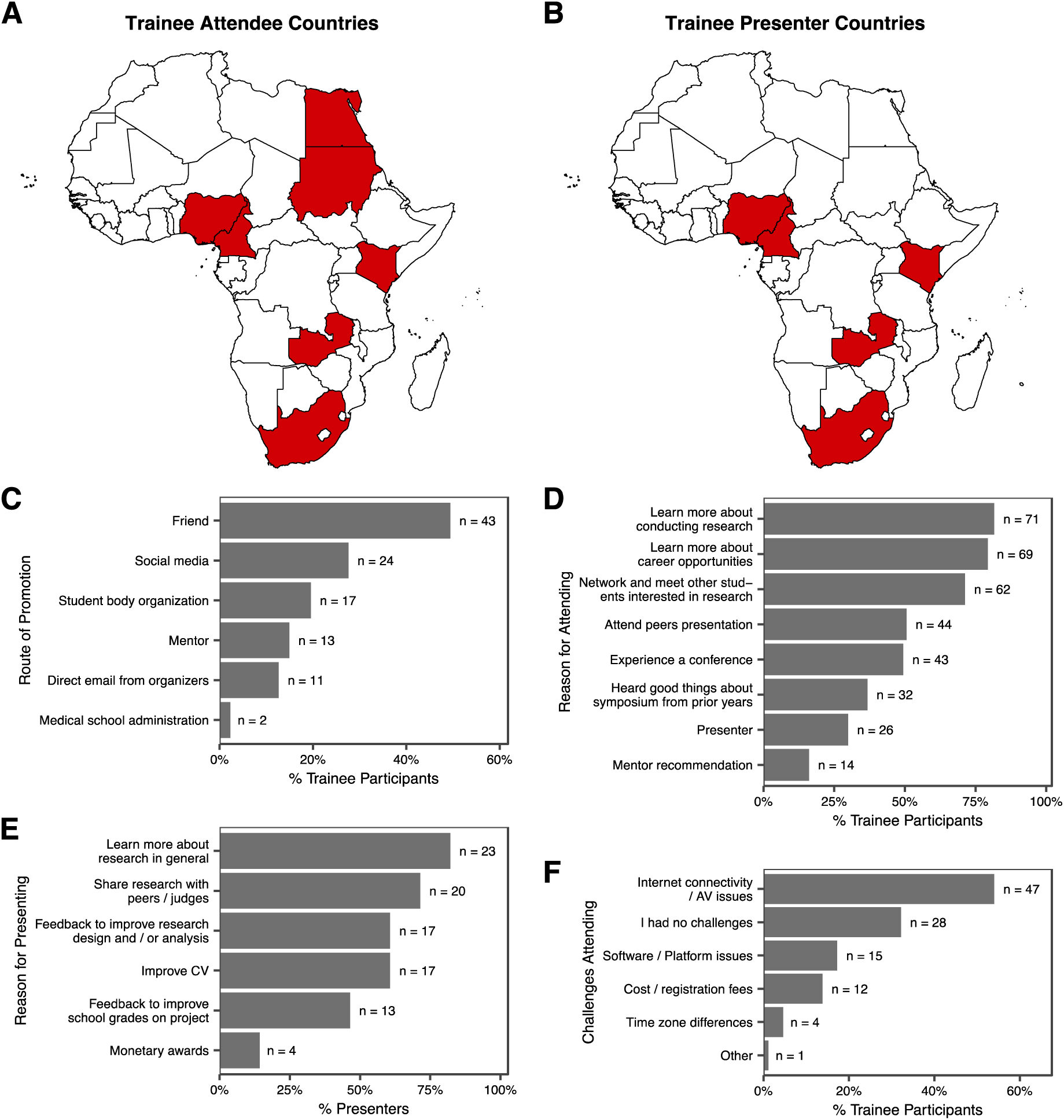
(A) Symposium trainee attendee and (B) trainee presenter home countries, shown in red. (C) Routes through which trainee attendees (n=87) reported learning of the symposium. (D) Trainee (n=87) reasons for attending. (E) Trainee presenter reasons for submitting abstracts (n=28). (F) Challenges trainees (n=87) faced in attending the symposium. In C-F, respondents were permitted to provide multiple answers to each question.

**Table 1.** Attendee and Presenter Baseline Characteristics.

|  | Number of Trainee Participants | Percent of Trainee Participants | Number of Non-Trainee Participants | Percent of Non-Trainee Participants |
| --- | --- | --- | --- | --- |
| **Responses of all Attendees** |  |  |  |  |
| **Gender** |  |  |  |  |
| Female | 44 | 50.6% | 4 | 57% |
| Male | 42 | 48.3% | 3 | 43% |
| Prefer not to say | 1 | 1.1% | 0 | 0% |
| Non-binary | 0 | 0% | 0 | 0% |
| **Occupation** |  |  |  |  |
| Medical Student | 76 | 87.4% | NA | NA |
| Junior Doctor (Intern) | 9 | 10.3% | NA | NA |
| Other student | 2 | 2.3% | NA | NA |
| Consultant | NA | NA | 3 | 43% |
| Other medical doctor | NA | NA | 2 | 29% |
| Other | NA | NA | 2 | 29% |
| **Method of attendance** |  |  |  |  |
| In-Person | 61 | 70.1% | 2 | 29% |
| Virtual | 26 | 29.9% | 5 | 71% |
| **Medical School Country** |  |  |  |  |
| Zambia | 72 | 82.8% | 4 | 57% |
| Egypt | 6 | 6.9% | 1 | 14% |
| Kenya | 4 | 4.6% | 0 | 0% |
| Nigeria | 2 | 2.3% | 1 | 14% |
| Cameroon | 1 | 1.1% | 0 | 0% |
| South Africa | 1 | 1.1% | 0 | 0% |
| Sudan | 1 | 1.1% | 0 | 0% |
| India | 0 | 0.0% | 1 | 14% |
| **Country of Residence** |  |  |  |  |
| Zambia | 72 | 82.8% | 5 | 71% |
| Egypt | 6 | 6.9% | 1 | 14% |
| Kenya | 4 | 4.6% | 0 | 0% |
| Nigeria | 2 | 2.3% | 1 | 14% |
| Cameroon | 1 | 1.1% | 0 | 0% |
| South Africa | 1 | 1.1% | 0 | 0% |
| Sudan | 1 | 1.1% | 0 | 0% |
| **Highest Educational Degree Completed** |  |  |  |  |
| None | 51 | 58.6% | 0 | 0% |
| Bachelor's Degree | 23 | 26.4% | 1 | 14% |
| MBBS/ MBChB / MD | 13 | 14.9% | 6 | 86% |
| **First member of family to have education beyond high school** |  |  |  |  |
| Yes | 27 | 31.0% | 1 | 14% |
| No | 60 | 69.0% | 6 | 86% |
| **First member of family to attend medical school** |  |  |  |  |
| Yes | 66 | 75.9% | 5 | 71% |
| No | 21 | 24.1% | 2 | 29% |
| **Monthly Household Income (USD)** |  |  |  |  |
| Less than 250 | 39 | 44.8% | 2 | 29% |
| 250 - 499 | 10 | 11.5% | 0 | 0% |
| 500 - 999 | 3 | 3.4% | 1 | 14% |
| 1000 - 2999 | 3 | 3.4% | 3 | 43% |
| Prefer not to say | 22 | 25.3% | 0 | 0% |
| (No Response) | 10 | 11.5% | 1 | 14% |
| **Member of family has conducted scientific or biomedical research** |  |  |  |  |
| No | 72 | 82.8% | 4 | 57% |
| Yes | 15 | 17.2% | 3 | 43% |
| **Have previously interacted with fellow students in a forum dedicated to presenting or discussing research** |  |  |  |  |
| No | 51 | 58.6% | 2 | 29% |
| Yes | 26 | 29.9% | 4 | 57% |
| Not applicable | 5 | 5.7% | 1 | 14% |
| (No Response) | 5 | 5.7% | 0 | 0% |
| **Have had formal training in research** |  |  |  |  |
| No | 53 | 60.9% | 0 | 0% |
| Yes | 34 | 39.1% | 7 | 100% |
| **Have previously attended a mentoring or scientific conference** |  |  |  |  |
| No | 45 | 51.7% | 1 | 14% |
| Yes | 42 | 48.3% | 6 | 86% |
| **Preferred Meeting Format** |  |  |  |  |
| In-Person | 58 | 66.7% | 4 | 57% |
| Hybrid | 23 | 26.4% | 3 | 43% |
| Virtual | 2 | 2.3% | 0 | 0% |
| (No Response) | 4 | 4.6% | 0 | 0% |
| **Responses of (Trainee) Presenters** |  |  |  |  |
| **Gender** |  |  |  |  |
| Female | 19 | 68% | N/A | N/A |
| Male | 9 | 32% | N/A | N/A |
| **Submitter Category** |  |  |  |  |
| Current Student | 19 | 68% | N/A | N/A |
| Recent Graduate | 9 | 32% | N/A | 32% |
| **Method of attendance** |  |  |  |  |
| In-Person | 15 | 54% | N/A | N/A |
| Virtual | 13 | 46% | N/A | N/A |
| **Medical School Country** |  |  |  |  |
| Zambia | 21 | 75% | N/A | N/A |
| Kenya | 3 | 11% | N/A | N/A |
| Cameroon | 2 | 7% | N/A | N/A |
| Nigeria | 1 | 4% | N/A | N/A |
| South Africa | 1 | 4% | N/A | N/A |
| **Highest Educational Degree Completed** |  |  |  |  |
| None | 12 | 43% | N/A | N/A |
| Bachelor's Degree | 7 | 25% | N/A | N/A |
| MBBS / MBChB / MD | 9 | 32% | N/A | N/A |
| **Have had formal training in research** |  |  |  |  |
| No | 12 | 43% | N/A | N/A |
| Yes | 16 | 57% | N/A | N/A |
| **Have previously attended a mentoring or scientific conference** |  |  |  |  |
| No | 15 | 54% | N/A | N/A |
| Yes | 13 | 46% | N/A | N/A |
| **Have previously written an abstract or presented at a research conference** |  |  |  |  |
| No | 13 | 46% | N/A | N/A |
| Yes | 15 | 54% | N/A | N/A |

Among trainees, 51 (58.6%) held no post-secondary academic degree, while 13 (14·9%) held medical degrees (table 1). Sixty-six (75·9%) were first in their families to attend medical school, and 27 (31·0%) were first in their families to pursue education beyond high school. Over half reported monthly household incomes below 500 USD. Most trainees had minimal prior research training or exposure, with 53 (58·6%) reporting no formal research education and 72 (82·8%) saying no family members had ever conducted scientific or biomedical research (table 1). Forty-five (51·7%) had never attended a scientific or mentoring conference.

### Trainee Presenter Characteristics

Twenty-eight trainees submitted abstracts and presented at the symposium. All 28 (100%) completed a supplemental presenter survey. The presenters gave 30 presentations (two presented multiple abstracts), of which 26 were research and 4 were case reports. Fifteen (54%) presented in-person.

Most presenters attended from Zambia (n=21, 75%), followed by Kenya (n=3, 11%), Cameroon (n=2, 7%), Nigeria (n=1, 4%) and South Africa (n=1, 4%; figure 1B). Two thirds were female (n=19, 68%; table 1). Most (n=19, 68%) were current medical students and many reported no formal education in research (n=12, 43%). Fifteen (54%) had never attended a conference and 13 (46%) had never previously written an abstract or presented at a conference. Eight of 11 (72·7%) prize recipients were female.

Abstracts covered diverse clinical and scientific subjects. Top-scoring research abstracts variously addressed cervical cancer surveillance, cancer treatment outcomes, hypertension monitoring in people with HIV, pediatric sickle cell disease management, renal dysfunction as a predictor of COVID-19 outcomes, epidemiology and quality-of-life impacts of chronic leg ulcers, epidemiology of neural tube defects, and medical student attitudes towards careers in neurology and neurosurgery. Top case reports included cases of pediatric malaria and small-bowel obstruction due to a retained surgical sponge (gossypiboma).

### Motivations and Barriers Affecting Symposium Attendance

Attendees learned of the symposium primarily through peer networks, with many citing multiple sources. Forty-three (49·4%), 24 (27·6%), and 17 (19·5%) of trainees respectively heard of the symposium from a friend, social media, and/or medical school student organizations (figure 1C). In comparison, just 13 (14·9%) heard through a mentor and just 2 (2·3%) from school administration.

Trainees reported multiple motivations for symposium attendance (figure 1D). Significant majorities hoped to gain insight into research methodologies (n=71, 81·6%), explore medical career opportunities (n=69, 79·3%), and/or establish connections and engage with fellow students interested in research (n=62, 71·3%). Half attended to gain conference experience (n=43, 49·4%) and/or support peers’ presentations (n=44, 50·6%). Thirty-two (36·8%) cited positive feedback from previous years as a motivation, while 14 (16·1%) said a mentor recommended they attend. Twenty-six (30%) reported attending to present their work.

The most common motivation for presenting (figure 1E) was desire to broaden understanding of research (n = 23, 82·1%), followed by desire to share findings with peers and judges (n=20, 71·4%) and receive feedback to refine project design and analysis (n=17, 60·7%). Seventeen (60·7%) wanted to improve their curriculum vitae, while 13 (46·4%) hoped for feedback to enhance project school grades. Monetary prizes were the least prevalent motivation, with only 4 (14·3%) citing them as a reason for presenting.

Trainees were also surveyed on barriers to attendance. Twenty-eight (32·2%) reported no difficulties, but 47 (54·0%) mentioned technical issues primarily attributed to poor network connections (network and audiovisual issues disrupted part of the symposium’s first day; figure 1F). Twelve (13·8%) said the registration fee of 5 to 10 USD posed a challenge, 15 (17·2%) struggled with software compatibility for virtual participation, 4 (4·6%) experienced challenges due to time zone differences, and 1 (1.1%) had difficulty securing time off.

### Trainee Satisfaction with Medical School Research Training

Many trainees reported deficiencies in research training and support at their home institutions. Only 46 (52·9%) somewhat agreed or strongly agreed that their medical schools encouraged students to pursue research during training, while just 47 (54·0%) somewhat or strongly agreed that their schools provided research opportunities (figure 2A). Only 34 (39·0%) somewhat or strongly agreed that their schools offered sufficient research training. Less than 5% (n=4) strongly agreed that they personally had adequate access to research mentorship through their school or that students at their school were satisfied with access to research training and mentorship.

**Figure 2.**
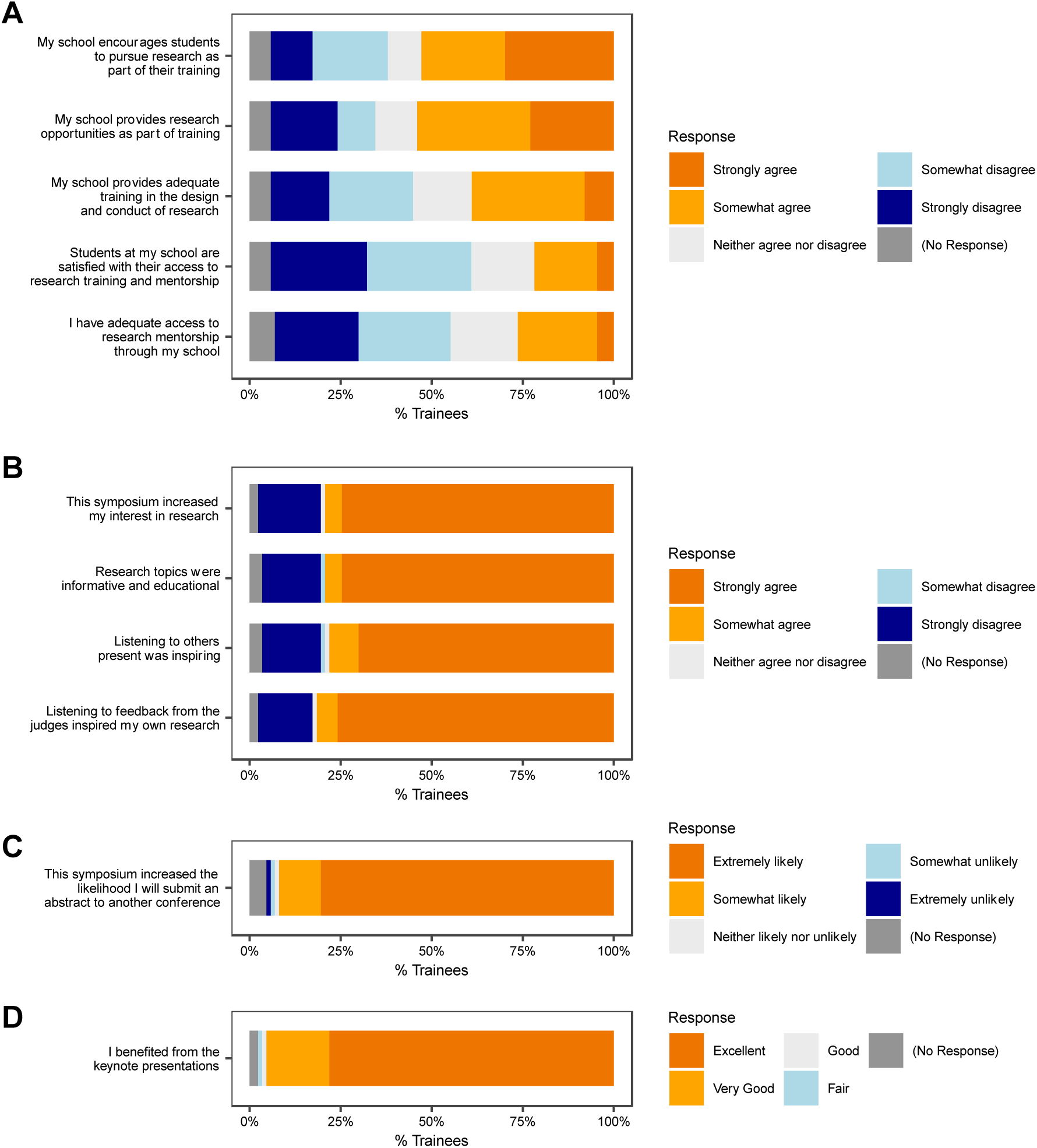
Satisfaction of trainees with research training and opportunities in medical school and self-reported benefits of attending the symposium. (A) Trainees’ (n=87) satisfaction with access to medical training, support, and mentorship at their home institutions. (B) Impact of the symposium on trainee interest in research. (C) Impact of the symposium on trainee interest in submitting to future conferences. (D) Trainees self-reported benefit from symposium keynote presentations. Data portrayed in the figure are also reported in tabular form in Supplementary Table 2 (ST2).

In a free-text response to whether medical schools encouraged research as part of training, a Zambian student wrote: “They encourage us to pursue research but do not offer the resources for it.” A South Sudanese national attending medical school in Egypt wrote: “We don’t have research opportunities in Egypt for foreign students.” A Zambian student described poor access to research mentorship, “because the supervisor has a lot of students to attend to,” while a second wrote, “I personally am interested in research but I don’t know who to go to for further guidance.”

### Benefits of the Symposium

Trainees reported substantial benefits from symposium attendance. Most agreed the symposium increased their interest in research (n=69, 79·3%), research topics were informative and educational (n=69, 79·3%), listening to others present was inspiring (n=68, 78·1%), and judge feedback inspired their own research (n=71, 81·6%; figure 2B). Eighty trainees (92%) said the symposium made them somewhat or extremely more likely to submit an abstract to a future conference (figure 2C), while 84 (96.6%) reported “strong”, “very strong”, or “excellent” benefit from keynote lectures (figure 2D). To understand which trainees most benefited from the symposium, we defined each question in figures 2B-D as a “benefit category”, then tallied numbers of categories in which each trainee reported benefit (see Methods). All trainees benefited in at least one category, with 60 (70·6%) benefitting in all six categories and another 10 (11·8%) benefitting in five categories (table 2). Notably, the degree of self-reported benefit remained consistent regardless of trainees’ gender, prior research training, satisfaction with institutional research support, prior conference experience, household income, or level of family education.

**Table 2.**
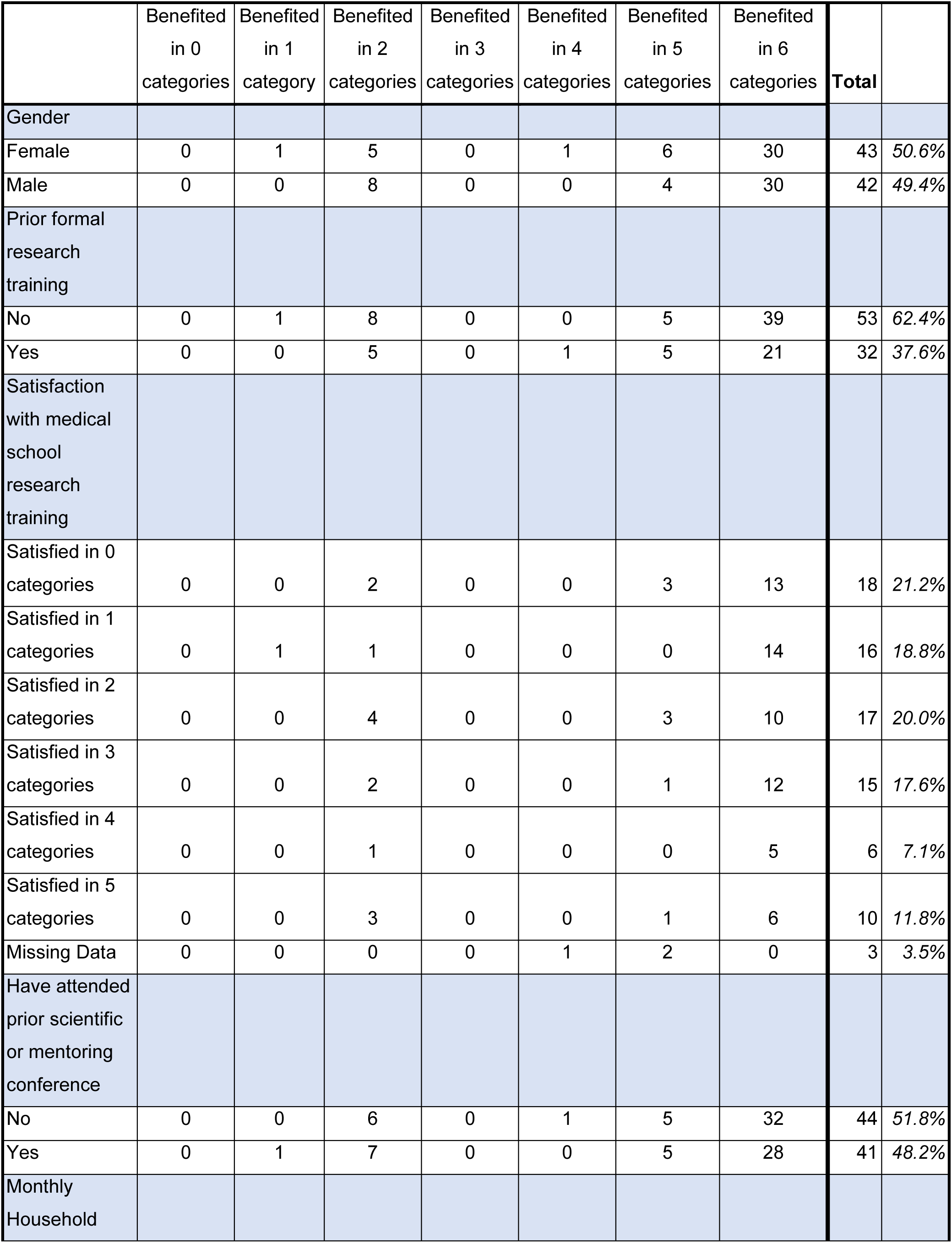
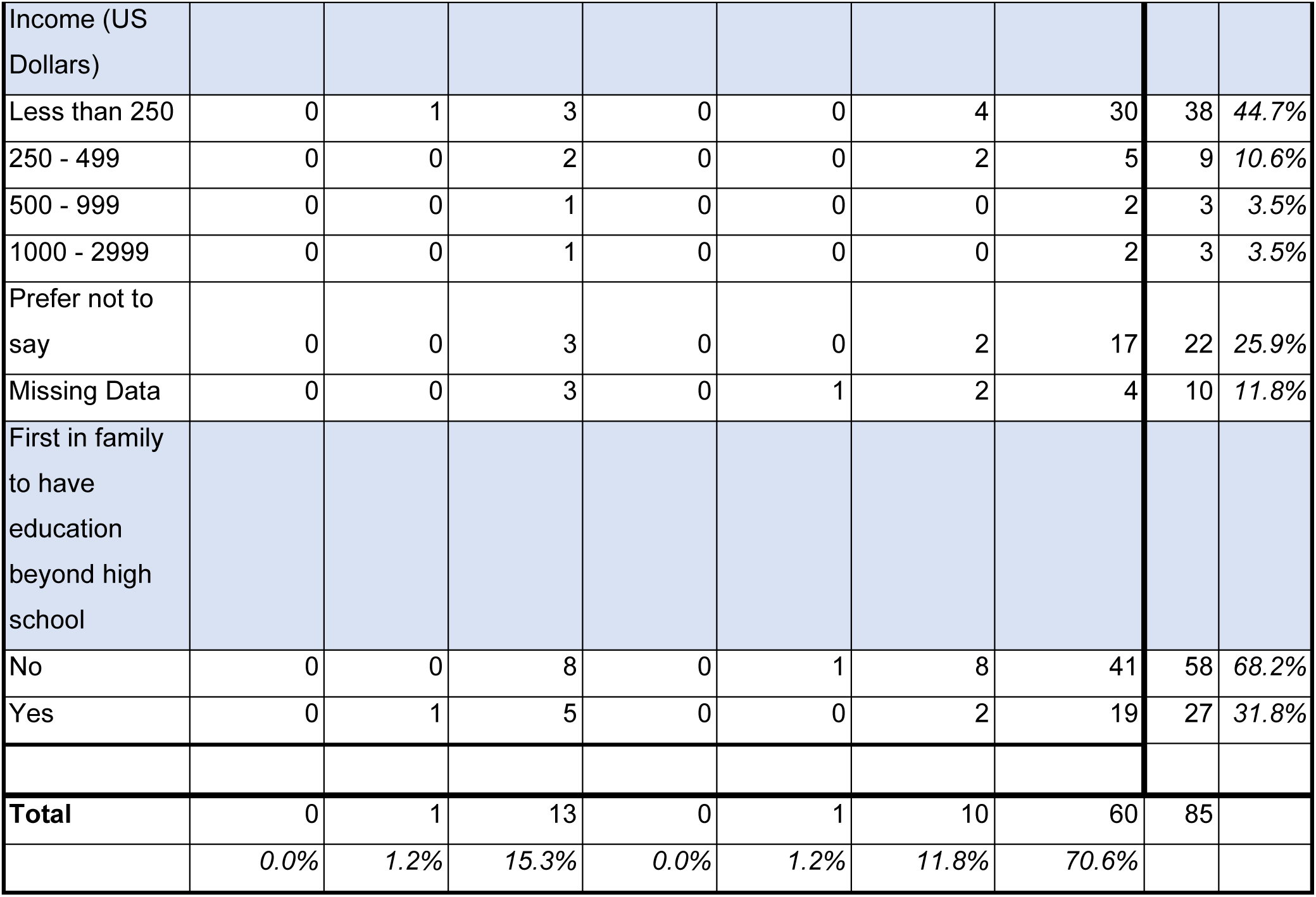
Number of categories in which trainees reported benefiting from the symposium, subdivided by trainee baseline characteristics.

Abstract presenters also reported strong benefits from presenting their work. Twenty-two (79%) said it was their first time receiving formative feedback from conference judges, while 24 (86%) said fellow attendees reached out individually in-person or electronically with feedback or questions after their presentations (figure 3A). Twenty-six (93%) said they gained new ideas to extend current research and 27 (96%) planned to submit their work to future conferences (figure 3A). All but one (96%) said it was “probably true” or “definitely true” that the symposium improved the quality of their work (figure 3B). Asked which aspects had improved, 24 (86%) cited writing and presentation, 18 (64%) interpretation of results, 16 (57%) data analysis, and 9 (32%) data collection (figure 3C).

**Figure 3.**
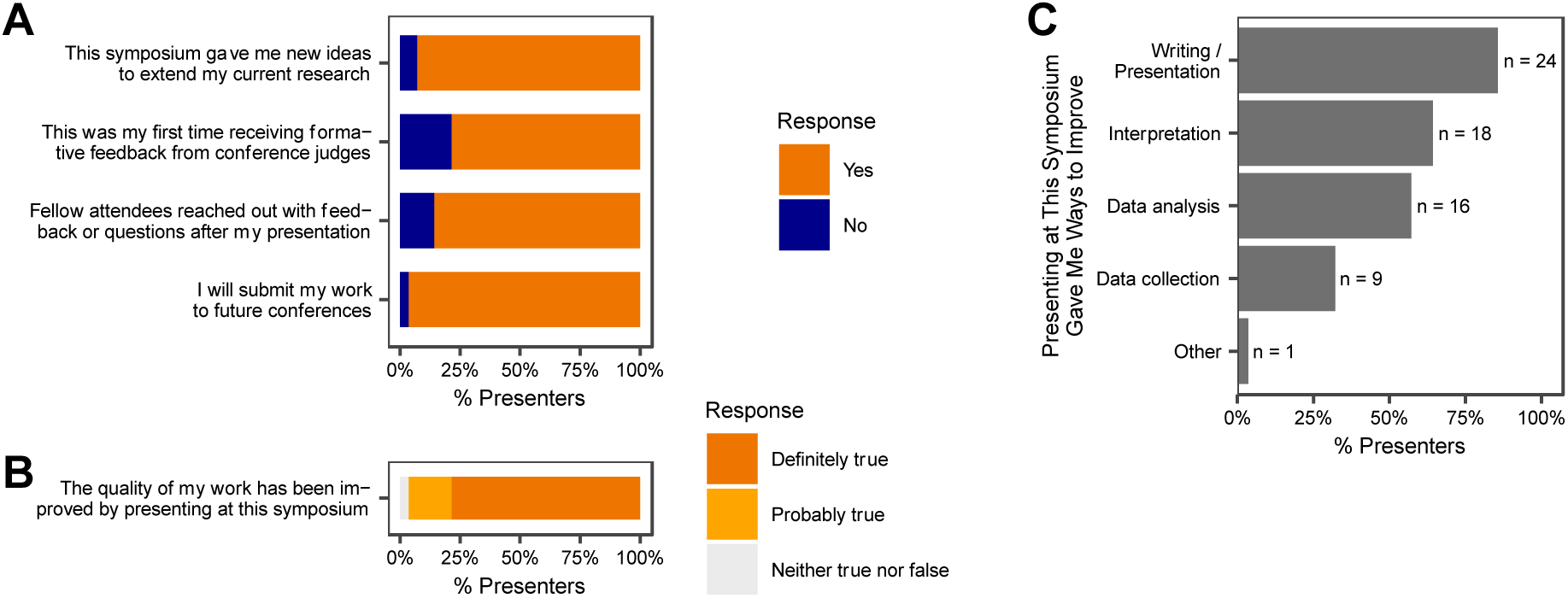
Self-reported benefits of the symposium for trainee abstract presenters (n=28) (A) Impact of the presenting experience on presenter research future directions, scientific networking and feedback, and desire to participate in future conferences. (B) Self-perceived benefit of the symposium on the quality of presenters’ work. (C) Self-perceived aspects of presenters’ projects that improved due to presenting at the symposium (multiple responses permitted). Data portrayed in the figure are also reported in tabular form in Supplementary Table 2 (ST3).

### Trainee Response Themes

To further explore which aspects of the symposium were most impactful, we performed thematic analysis of free-text survey responses. In response to the question “What did you like the most about the symposium,” the largest fraction cited trainee presentations. One student wrote, “Watching my peers present their research was so inspirational because [it] made me realize that as a student I could conduct research […] myself.” Another liked “The fact that I got to present. It’s the best experience of anything I have had this year, honestly!” Keynote speakers, including one on trainee mental health, were praised. Attendees also highlighted the symposium’s educational aspects, networking opportunities, judge feedback, mentorship, and international scope. A student wrote “It exposed me to view research as being more of a necessity to [medical] advancement, than a must do,” while another liked that the symposium, “Provides a platform for researchers to showcase their findings on [an] international stage [and] share ideas with peers from different schools that otherwise would not cross paths.”

Responses to the questions “What did you like/dislike about the judging? What would you change about the judging?” highlighted consistent themes. Both presenters and non-presenters mentioned objectivity and honesty of judge evaluations, fair assessment of presenters, and benefit to attendees from judges’ feedback. One student wrote, “I liked that the judges did not only ask questions about the presentations but also gave guidance where needed on how to improve research taking and presentation,” while a second wrote, “I liked how the judges always pointed out the strengths in presenters and always made recommendations in an educative way.” A junior doctor commented, “The feedback was constructive and made me feel there is more room for improvement in my presentation and research moving forward,” and a student wrote, “The judging enlightened me and gave me motivation to improve my own research.” The commonest critique concerned limited time allotted for judges to comment.

When asked what they liked least or would change about the symposium, many said no changes were needed. Others mentioned technical, network, or logistical issues, issues with the venue, adding time and breaks to the program, improved interaction for virtual participants, facilitation of direct trainee-mentor discussions, and mechanisms to formally link mentees to mentors. Many recommended better publicizing future symposia, with one student writing, “next time it should be well advertised and invite as many people as possible [because] the meeting was so impactful.” Requests for future topics encompassed four main themes: 1) non-communicable diseases (hypertension, diabetes, neuroscience, mental health, and menstrual disorders), 2) tropical/infectious diseases, 3) research (funding, conduct, and promotion), and 4) career guidance (entrepreneurship in medicine, women in medicine, biomedical sciences, scholarships, and personal development).

## Discussion

Research exposure is vital to medical education^1,2^, but many African medical students lack access to robust research training and opportunities, threatening global health progress.^7,11^ Surveys at African medical schools have found that students express strong interest in conducting research, but say they face important barriers including absence of mentorship, methodologic knowledge, funding, and collaborations.^5,8,9^ We report results of a volunteer-organized research and mentoring symposium that attracted students and junior doctors from diverse medical schools across 7 African countries. Most attendees were first in their families to attend medical school, reported low household income, had no research background, had never attended a conference, and faced key deficiencies in research opportunities, training, and mentorship at their home institutions. The vast majority reported substantial benefits from the symposium, including increased knowledge of research, improvements to their projects based on feedback received, networking with scientifically inclined peers, and inspiration to initiate new research. These results extend findings from high-income settings showing that conferences can help students expand knowledge, pursue international projects, and grow professionally.^17,12^

The symposium had a number of features that enhanced its impact. The first involved formative feedback. Most attendees had never previously interacted with peers in a research forum where projects were scientifically critiqued, echoing findings from a survey at Ugandan medical schools in which students complained that their classes did not teach them to apply statistical and methodologic principles to real-world research.^8^ Presenters at our symposium received both written reviews and live formative feedback from judges in front of the full audience. In contrast to feedback methods that provide individual written comments to conference presenters,^18,19^ this format allowed all attendees to witness a process of scientific review and critique. Judges emphasized a “work-in-progress” mentality, suggesting ways of improving study design, analysis, and communication to prepare projects for publication or submission to field-specific conferences. Judges also highlighted presentation strengths as teaching points for fellow attendees. Both presenters and non-presenters praised the format, saying it enhanced research understanding and inspired them to improve current work and initiate new studies. Enthusiasm was high regardless of trainees’ demographics, prior research training, or level of satisfaction with institutional support. In one student’s words, “I liked the frankness [of the judges] in guiding the presenters…wherever they were [supposed] to make changes. That promotes change and development for young emerging medical researchers.” Indeed, the major complaint about the judging was that time limitations prevented even more detailed feedback.

Another key feature of the symposium involved gender balance. Gender inequality in medicine and research remains prevalent (including in Africa), reflected by metrics such as underrepresentation of female researchers in authorship.^20^ This often extends to female underrepresentation at conferences, which can discourage aspiring female medical researchers and deprive them of role models.^21^ Our symposium featured robust female representation. Although the judges were male, attendees were gender-balanced, all session moderators were women, and majorities of symposium organizers, keynote speakers, abstract presenters, and prize winners were female. Female attendees reported benefitting from the symposium in equal degree to males. The dynamic was encapsulated by a student who wrote that her favorite aspect was, “Learning more about research and the talks that helped with self confidence and how to go about life in medicine. Especially as a female, most presenters were female and that was encouraging to see because it shows me it can be done and I’m not being too ambitious for wanting to go out and do more and be more. If they could do it, I can do it too.”

Our study does have limitations. Attendees were self-selected and may not be representative of all African medical trainees. Not all attendees completed the survey, potentially biasing results. The study lacks long-term follow-up of participant outcomes. The limited budget (<10,000 USD) and reliance on volunteers hampered symposium implementation by impairing network stability, audiovisual support, and logistics. Funding limitations prevented award of travel scholarships, forcing most non-Zambian attendees to participate virtually. Finally, sparse participation by senior experts prevented pairing many aspiring researchers with long-term mentors.

This study highlights limitations in institutional research training, mentorship, and opportunities for African medical trainees, while demonstrating benefits of a trainee-focused research and mentorship symposium. The symposium provided students otherwise lacking research support with opportunities to present, receive and witness scientific feedback, develop scientific abilities, and network with peers and mentors. The symposium was organized by unpaid volunteers – most of them current African medical students or recent graduates – highlighting the enthusiasm among trainees for research exposure and the positive impacts achievable with even limited resources. Future symposia and other initiatives to support African medical trainee research can help inspire and develop new generations of research leaders and promote ongoing global health advances by and for Africa.

## Contributors

AWA, SMB, and AAN conceived of and designed the study. AAN, SMB, and AWA designed the survey. AWA and MMu obtained ethics approval. AAN, AWA, SMB, MMu, and MMa coordinated data collection. SMB and AAN conducted quantitative data analysis with input from all authors. MMa conducted thematic analysis with input from SMB and TK. AAN and SMB led the drafting and organization of the manuscript with contributions from MMu, MMa, TK and SJM. AWA, AK, and SM provided review and editing. AWA and SMB provided overall project supervision. All authors provided critical feedback and approved the final version of the manuscript.

## Data sharing

Data will be made available upon reasonable request after removal of all potentially identifying information.

## Declaration of interests

All authors declare no competing interests.

## Data Availability

All data produced in the present study are available upon reasonable request to the authors

## Acknowledgments

The authors would like to acknowledge Dr Tepwanji Mpetemoya for her work at the initial stages of the project, as well as symposium co-organizers, participants, donors, and abstract reviewers.

## Supplementary Materials

Supplementary Table ST1. Keynote Presenters at the 2022 Symposium

Supplementary Table ST2. Data associated with the satisfaction of trainees with research training and opportunities in medical school and self-reported benefits of attending the symposium.

Supplementary Table ST3. Self-reported benefits of the symposium for trainee abstract presenters

## Notes

### Competing Interest Statement

The authors have declared no competing interest.

### Funding Statement

This study did not receive any funding

### Author Declarations

Ethics approval was obtained from the Tropical Diseases Research Center (TDRC) Ethics Committee [TDREC/023/022] in Ndola, Zambia.

### Summary of Updates

Author name updated from Avis A to Anya A to align with authors other publications.

