## Supplementary Tables for "Educational Impact of a Research and Mentoring Symposium Emphasizing Formative Feedback for Medical Students and Early Career Doctors in Africa"

Supplementary Table ST1. Keynote Presenters at the 2022 Symposium

| Keynote Title | Speaker | Professional Title and Affiliation | Country |
|---|---|---|---|
| **The State of Health Research in Africa: Current Challenges, Solutions, and Opportunities** | Dr. Patricia Shinondo, MD | Pediatric Surgeon, Levy Mwanawasa University Teaching Hospital, Lusaka, Zambia; Vice President, Zambia Medical Association | Zambia |
| **Mentorship and Mentorship Opportunities - Choosing a Mentor** | Professor Nazik Hammad, MD, MSc, MEHP, FACP | Professor, Division of Medical Oncology, Queen's University, Kingston Health Science Center, Kingston, Ontario, Canada | Canada |
| **Translating Research into Policy Advancements** | Professor Ruanne Barnabas, MD | Chief, Division of Infectious Diseases, Massachusetts General Hospital, Boston, Massachusetts, USA; Professor of Medicine, Harvard Medical School, Boston, Massachusetts, USA; Professor of Epidemiology, Harvard T.H. Chan School of Public Health, Boston, Massachusetts, USA | USA |
| **Getting Started in Private Practice - The Do's and Don'ts'** | Dr. Emilia Marsden, MBChB, MMed Paeds | Pediatrician; Managing Director and Board Member, Pendleton Family Practice, Lusaka, Zambia | Zambia |
| **Advancing Gender Equity in Academia: Challenges, Strategies, and Institutional Change** | Dr. Miriam Mutebi, MD, MMed Surg, MSc | AORTIC President-Elect; Consultant Breast Surgical Oncologist and Assistant Professor, Aga Khan University Hospital, Nairobi, Kenya | Kenya |
| **Mental Health in Medical School** | Dr. Naeem Dalal, MD, BScHB, MBChB, MMed Psychiatry | Psychiatrist, University Teaching Hospital, Lusaka, Zambia; Public Health Chairperson, Zambia Medical Association | Zambia |
| **The Power of You - Identifying your Strengths and Getting Started in Achieving your Goals** | Olive Strachan, MBE | CEO, Olive Strachan Consultancy | United Kingdom; Dominica |

Supplementary Table ST2. Data associated with the satisfaction of trainees with research training and opportunities in medical school and self-reported benefits of attending the symposium.

(A) Trainees' (n=87) satisfaction with access to medical training, support, and mentorship at their home institutions.

| Question | (No Response) | Strongly disagree | Somewhat disagree | Neither agree nor disagree | Somewhat agree | Strongly agree |
|---|---|---|---|---|---|---|
| **My school encourages students to pursue research as part of their training** | 5 (5.7%) | 10 (11.5%) | 18 (20.7%) | 8 (9.2%) | 20 (23%) | 26 (29.9%) |
| **My school provides research opportunities as part of training** | 5 (5.7%) | 16 (18.4%) | 9 (10.3%) | 10 (11.5%) | 27 (31%) | 20 (23%) |
| **My school provides adequate training in the design and conduct of research** | 5 (5.7%) | 14 (16.1%) | 20 (23%) | 14 (16.1%) | 27 (31%) | 7 (8%) |
| **Students at my school are satisfied with their access to research training and mentorship** | 5 (5.7%) | 23 (26.4%) | 25 (28.7%) | 15 (17.2%) | 15 (17.2%) | 4 (4.6%) |
| **I have adequate access to research mentorship through my school** | 6 (6.9%) | 20 (23%) | 22 (25.3%) | 16 (18.4%) | 19 (21.8%) | 4 (4.6%) |

(B) Impact of the symposium on trainee interest in research.

| Question | (No Response) | Strongly disagree | Somewhat disagree | Neither agree nor disagree | Somewhat agree | Strongly agree |
|---|---|---|---|---|---|---|
| **This symposium increased my interest in research** | 2 (2.3%) | 15 (17.2%) | 0 (0%) | 1 (1.1%) | 4 (4.6%) | 65 (74.7%) |
| **Research topics were informative and educational** | 3 (3.4%) | 14 (16.1%) | 1 (1.1%) | 0 (0%) | 4 (4.6%) | 65 (74.7%) |
| **Listening to others present was inspiring** | 3 (3.4%) | 14 (16.1%) | 1 (1.1%) | 1 (1.1%) | 7 (8%) | 61 (70.1%) |
| **Listening to feedback from the judges inspired my own research** | 2 (2.3%) | 13 (14.9%) | 0 (0%) | 1 (1.1%) | 5 (5.7%) | 66 (75.9%) |

(C) Impact of the symposium on trainee interest in submitting to future conferences.

| Question | (No Response) | Extremely unlikely | Somewhat unlikely | Neither likely nor unlikely | Somewhat likely | Extremely likely |
|---|---|---|---|---|---|---|
| **This symposium increased the likelihood I will submit an abstract to another conference** | 4 (4.6%) | 1 (1.1%) | 1 (1.1%) | 1 (1.1%) | 10 (11.5%) | 70 (80.5%) |

(D) Trainees self-reported benefit from symposium keynote presentations.

| Question | (No Response) | Poor | Fair | Good | Very Good | Excellent |
|---|---|---|---|---|---|---|
| **I benefited from the keynote presentations** | 2 (2.3%) | 0 (0%) | 1 (1.1%) | 1 (1.1%) | 15 (17.2%) | 68 (78.2%) |

Supplementary Table ST3. Self-reported benefits of the symposium for trainee abstract presenters (n=28)

(A) Impact of the presenting experience on presenter research future directions, scientific networking and feedback, and desire to participate in future conferences.

| Question | No | Yes | |
|---|---|---|---|
| **This symposium gave me new ideas to extend my current research** | 2 | 26 | 92.86% |
| **This was my first time receiving formative feedback from conference judges** | 6 | 22 | 78.57% |
| **Fellow attendees reached out with feedback or questions after my presentation** | 4 | 24 | 85.71% |
| **I will submit my work to future conferences** | 1 | 27 | 96.43% |

(B) Self-perceived benefit of the symposium on the quality of presenters' work.

| Question | Neither true nor false | Probably true | Definitely true | |
|---|---|---|---|---|
| **The quality of my work has been improved by presenting at this symposium** | 1 | 5 | 22 | 78.57% |

(C) Self-perceived aspects of presenters' projects that improved due to presenting at the symposium (multiple responses permitted).

| Presenting at This Symposium Gave Me Ways to Improve | Total Participants | Number of Presenters | % Presenters |
|---|---|---|---|
| **Writing / Presentation** | 28 | 24 | 86% |
| **Interpretation** | 28 | 18 | 64% |
| **Data analysis** | 28 | 16 | 57% |
| **Data collection** | 28 | 9 | 32% |
| **Other** | 28 | 1 | 4% |
